# Sensing of COVID-19 Antibodies in Seconds via Aerosol Jet Printed Three Dimensional Electrodes

**DOI:** 10.1101/2020.09.13.20193722

**Authors:** Md. Azahar Ali, Chunshan Hu, Sanjida Jahan, Bin Yuan, Mohammad Sadeq Saleh, Enguo Ju, Shou-Jiang Gao, Rahul Panat

## Abstract

*Rapid diagnosis is critical for the treatment and prevention of diseases. In this research, we report sensing of antibodies specific to SARS-CoV-2 virus in seconds via an electrochemical platform consisting of gold micropillar array electrodes decorated with reduced graphene oxide and functionalized with recombinant viral antigens. The array electrodes are fabricated by Aerosol Jet (AJ) nanoparticle 3D printing, where gold nanoparticles (3-5nm) are assembled in 3D space, sintered, and integrated with a microfluidic device. The device is shown to detect antibodies to SARS-CoV-2 spike S1 protein and its receptor-binding-domain (RBD) at concentrations down to 1pM via electrochemical impedance spectroscopy and read by a smartphone-based user interface. In addition, the sensor can be regenerated within a minute by introducing a low-pH chemistry that elutes the antibodies from the antigens, allowing successive testing of multiple antibody samples using the same sensor. The detection time for the two antibodies tested in this work is 11.5 seconds. S1 protein sensing of its antibodies is specific, which cross-reacts neither with other antibodies nor with proteins such as Nucleocapsid antibody and Interleukin-6 protein. The proposed sensing platform is generic and can also be used for the rapid detection of biomarkers for other infectious agents such as Ebola, HIV, and Zika, which will benefit the public health*.

## Main

Pandemics and epidemics caused by emerging infectious agents such as SARS-CoV, SARS-CoV-2, influenza viruses, ZIKA virus, and Ebola virus have seriously affected human health, and led to lost economic activity and challenged health-care systems.^1-3^ Early detection of the infection allows isolation of the patients and contact tracing that leads to saving of lives and resuming of economic activity. Even after opening of the economy, secondary spikes are likely to occur, requiring low-cost, rapid methods for early detection of the infection. Recently, the SARS-CoV-2 virus has caused the COVID-19 pandemic with a significant global impact.^4-6^ A testing platform that can detect the infection of this virus within seconds can be of significant help in saving lives. This is especially true since a significant proportion of the patients transmitting the disease are asymptomatic.^7^ In addition, such a device will open up sectors of the economy such as educational institutions, theme parks, airlines, and cruise ships where large gatherings of people are inevitable.

Two primary approaches have been used to detect the COVID-19 infection. The first approach utilizes the genetic sequence of the virus and includes genomic sequencing, CRISPR-based test^8^, and reverse transcription real time quantitative PCR (RT-qPCR).^8-10^ These methods are relatively accurate but are generally slow because of multiple sample processing steps and the need to ship the sample to a testing laboratory. The accuracy of these methods is also dependent upon the protocol of sampling, preservation, transportation, and processing of the samples.^11^ Since the viral RNA polymerase does not possess the proof-reading function, it leads to a high mutation rate. As a result, the accuracy of these methods is compromised if the virus mutates in the targeting genomic region, which occurs frequently.^12^ The second approach is to detect specific antibodies to the viral antigens.^13^ These serological methods include enzyme-linked immunosorbent assay (ELISA) and lateral flow immunoassay, some of which often give on-site results within ten minutes without requiring sophisticated sample preparation.^14, 15^ Current assays for detection of COVID-19 infections are based on structural proteins including spike protein (S-protein) and/or nucleocapsid protein (N-protein).^16^ These assays have a relatively low sensitivity, a false positive rate of about 5-11%,^13^ and usually a long sample preparation time of at least 0.5-2 hr. Hence, there is an urgent need to develop a sensitive and specific assay for rapid detection of SARS-CoV-2 infection within minutes or if possible, seconds; ideally within days of infection, which could particularly be useful for medically underserved areas if the readout is enabled by a smartphone. Electrochemical sensing is a promising method for the detection of pathogens or antibodies.^17, 18^

In this method, the formation of antibody-antigen complex is detected via electrochemical transduction. The sensitivity, specificity, and speed of detection, however, will depend upon the electrochemical cell and the geometry and surface chemistry of the electrodes, in addition to the antigen and assay procedures. In conventional electrochemical sensors, the electrodes are typically planar 2D structures which are often decorated with nanomaterials to increase their surface area.^19^

We note that biosensing devices have evolved with the developments in microelectronics – moving from bio-MEMS to miniaturized lab-on-chip constructs made by advanced photolithographic techniques.^20^ Recently, nanoparticle-based 3D printing methods have emerged in microelectronics that enable intricate geometries, material combinations, and custom microstructures. These methods offer many exciting advantages over conventional lithography such as a simple two-step fabrication process controlled by CAD programs, customizability, and prototypability. Amongst the different additive manufacturing methods, Aerosol Jet (AJ) nanoparticle 3D printing is a technique that uses a stream of aerosolized droplets to deposit an array of functional materials at a resolution of 10 μm and has been used to fabricate various electronic devices.^21^ Until recently, AJ printing was primarily used to create 2D planar structures as electronic components in devices.^22^ Recently, however, Saleh *et al.^23^* have used the dynamics of the aerosol microdroplets to create 3D device geometries with aspect ratios as high as 50:1, providing pathways to create complex and intricate geometries with high surface-to-volume ratios for electrodes.

The work presented in this paper was thus motivated by two factors. First, we aimed to utilize the latest advance in microelectronics fabrication, namely nanoparticle 3D printing, to solve the pressing need of developing a rapid test for the detection of antibodies to COVID-19 infection. The focus was to use AJ nanoparticle 3D printing to create three-dimensional electrode geometry and develop a unique surface chemistry that significantly enhances the transport of diffusing species in an electrochemical cell. The second aim was to use electrochemical transduction to detect COVID-19 antibodies within seconds and with a regeneration capability, along with a readout enabled by a smartphone-based platform. The focus was to develop a chemistry that can regenerate the device within minutes and establish selectivity, sensitivity, reproducibility, and repeatability of this sensor platform. We also discuss the significant positive implications of this work to the benefit of public health.

## Results

### Design and Construction of COVID-19 Test Chip with 3D Electrodes

We first describe the 3D Printed COVID-19 Test Chip (3DcC) platform. The schematic of the 3DcC device along with AJ printing of the three-dimensional electrodes is shown in Fig. 1. The functionalization of 3D electrodes by viral antigens is also a critical step in 3DcC device formation and is described in Fig. 2. Although details of the fabrication process are given in Materials and Methods, the device construction and surface chemistry are central to the function of the sensor and hence briefly described below. Fig. 1a shows a glass slide coated with patterned chromium (5 nm thick) and gold (100 nm thick) which formed the base layer of the three electrodes (working electrode or WE, counter electrode or CE, and reference electrode or RE) of an electrochemical cell. Fig. 1b shows the AJ printing process of the micropillar array on the WE shown in Fig. 1a. The AJ printer breaks the gold nanoparticle ink in the vial via ultrasonication into microdroplets, each containing the gold nanoparticles (2-5 nm in diameter). The microdroplets were carried to the nozzle via an inert gas (N**2**) and aerodynamically focused on the WE, which was heated to 150 °C. Figs. 1c and 1d show a CAD (Computer Aided Design) controlled process of droplet dispense to form individual pillars. The deposition was in the form of micro-rings of gold nanoparticle ink. Once one layer was printed, the solvent evaporated due to the heat from the substrate, forming solidified ‘dry’ material containing the nanoparticles and binders. When the next ring was printed, the surface tension of the solvent in the ring allowed the micropillars to be built without the use of any support structures. Each layer of the rings was about 5-10 μm thick and formed within a fraction of a second. A succession of these processes formed a 10×10 micropillar array consisting of unsintered nanoparticles and binders. Upon sintering of the printed structure, the gold micropillars for the electrode were formed. Fig. 1e shows the schematic of the construction of the PDMS housing by replica-molding method, which involved using polymethylmethacrylate (PMMA) mold and a polydimethylsiloxane (PDMS) mold to create a PDMS microfluidic channel. The RE, shown in Fig. 1a was coated with a thin silver/silver chloride (Ag/AgCl) layer via a shadow mask. The PDMS housing was then placed manually on the glass slide containing the micropillar array electrode and other electrodes (CE and RE). The complete construction of the 3DcC device is shown in Fig. 1f. Note that fluid could be introduced in the device by tubes inserted into the microfluidic channel.

**Figure 1.**
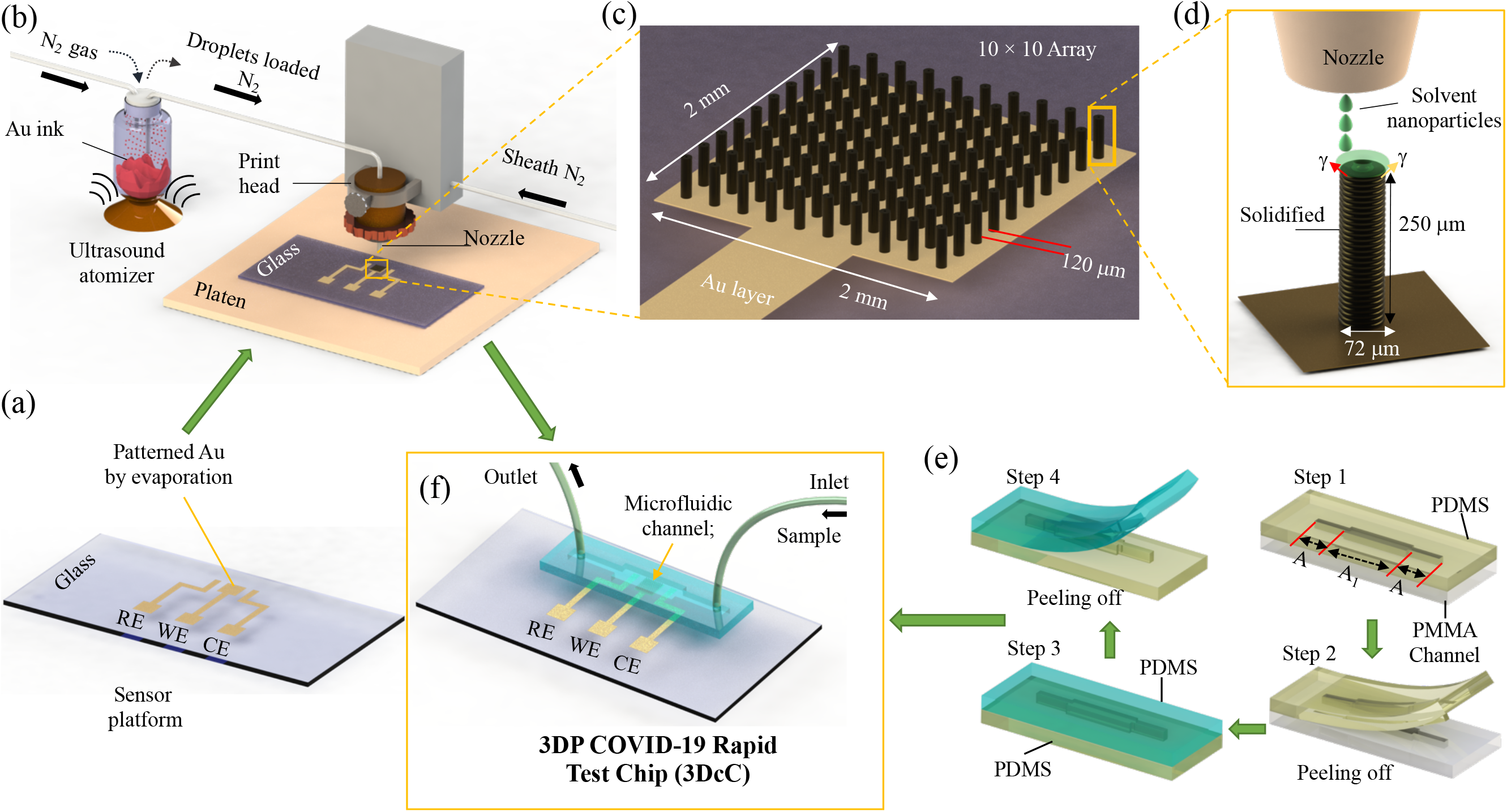
Schematic of the Manufacturing process of the 3D Printed COVID-19 Test Chip (3DcC) by Aerosol Jet nanoparticle 3D printing. (a) Glass substrate with patterned gold film forming the base for working electrode (WE), counter electrode (CE) and reference electrode (RE) of the electrochemical cell of 3DcC. (b) Construction of Aerosol Jet machine where gold ink is converted into an aerosol consisting of micro-droplets using ultrasonic energy and transported to a nozzle by N_2_ gas where it is focused on the gold film of the WE using a sheath gas (also N_2_) to form the micropillars. The entire process is digitized with the CAD program controlling the printing process. (c) An AJ printed 10×10 gold micropillar array where pillar-to-pillar gap is indicated. (d) Details of AJ printing of a single micropillar where rapid layer-by-layer stacking of the micro-rings of the nanoparticle ink are achieved using surface tension (g) of the printed ink. The entire process is achieved without the use of any support structure. Once a layer was printed, the ink loses solvents due to the heat from the platen (which was heated to 150 °C using a custom heater). The dried ink provides a base to receive the next micro-ring; and the process is repeated. (e) Process of fabrication of the PDMS housing of the 3DcC device. A PDMS structure is created using a PMMA master mold using replica molding. This structure then acts as a mold for the PDMS housing that contains a cavity for microfluidic channel as shown. (f) The 3DcC device formed by placing the PDMS housing with microfluidic channel on the glass substrate with the micropillar electrodes. Prior to this step, the micropillar electrodes were functionalized with reduced graphene oxide (rGO) and viral antigens as described in Fig. 2.

An important step in the working of the 3DcC device is the functionalization of the 3D printed microelectrode by viral antigens for detection of antibodies from the fluid introduced in the electrochemical cell. This process is depicted in Figs. 2a-2d. The bare 3D micropillar electrode is shown in Fig. 2a. The micropillar electrode array functionalized using reduced graphene oxide (rGO) is shown in Fig. 2b. The carboxylated (-COOH) groups of rGO are also shown in the Fig. 2b. Due to the π-π interactions amongst the coated rGO, the coating was expected to be non-uniform.^24^ Fig. 2c shows antigens bonded with the rGO flakes of the electrode. This was achieved by activating the -COOH groups of rGO using a coupling chemistry of EDC:NHS (1-Ethyl-3-(3-dimethylaminopropyl) carbodiimide, N-Hydroxy-Succinimide).^25^ This chemistry facilitates the formation of C-N co-valent bonding between rGO and antigens via an amidation reaction.^25^ Specifically, this reaction involved the EDC molecules acting as cross-linkers between -COOH and -NH**2** (i.e., amine group of the viral antigens) and NHS molecules acting as stabilizers during this reaction.^25^ Two SARS-CoV-2 viral antigens, namely, spike S1 and receptor binding domain (RBD), were separately used in this process (in different sensors). Schematic in Fig. 2d shows antibodies selectively binding with the antigens when a fluid containing antibodies was introduced in the chamber. It is noted that a treatment with bovine serum albumin (BSA) molecules was used to block non-specific sites of the sensor. The details of the electrode surface functionalization are described in Materials and Methods section.

**Figure 2.**
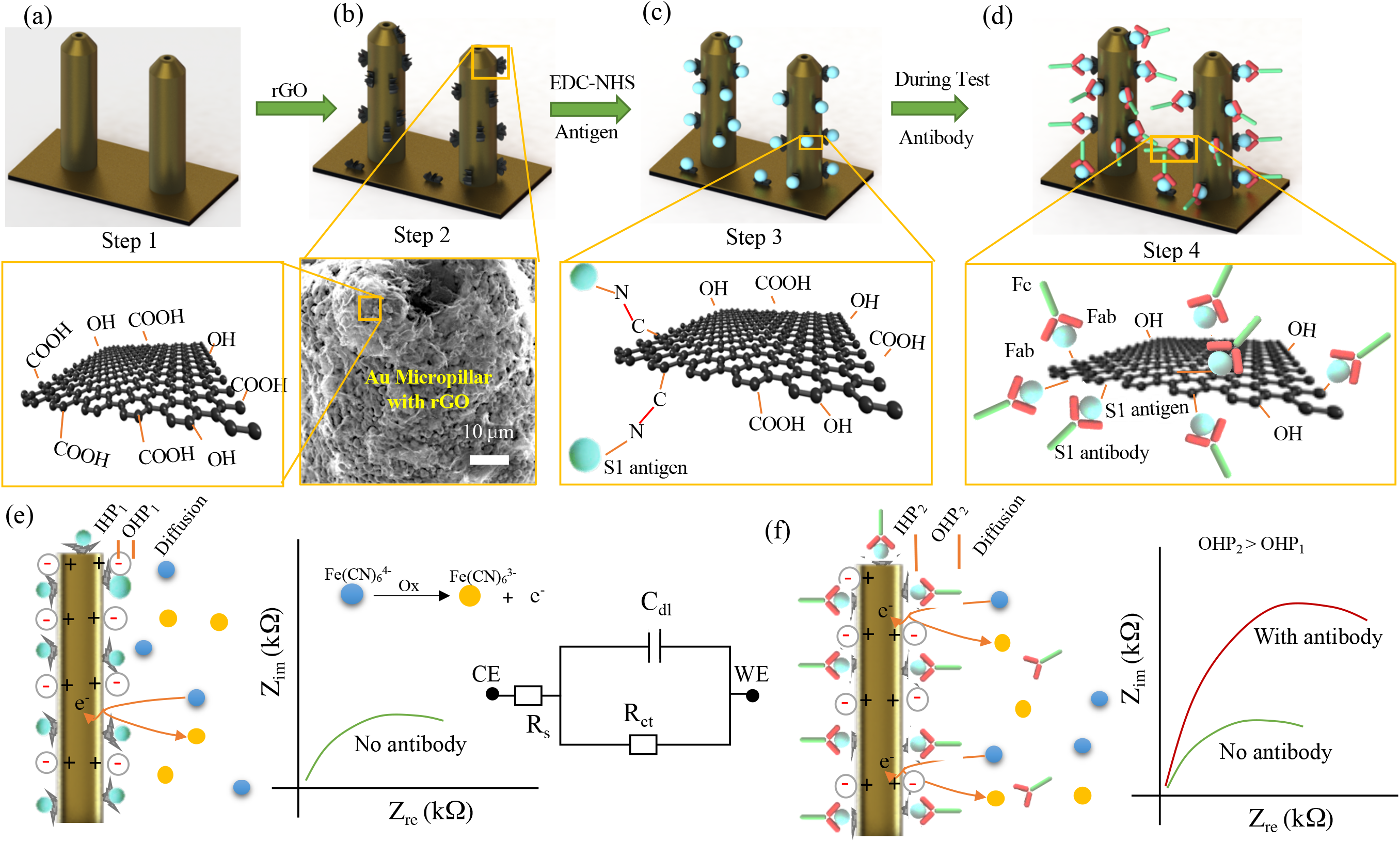
Functionalization of 3D Printed Micropillar Electrode and 3DcC Sensor Operation. (a) AJ printed gold micropillars prior to the surface treatment (step 1). (b) Coating of the electrodes by carboxylated (-COOH) reduced-graphene oxide (rGO) sheets by a simple drop-casting process (step 2). An SEM image shows the decoration of rGO sheets on the gold pillar. The electrostatic or *Van der Waals* interactions allow the rGO sheets to be connected to the micropillar. The surface porosity of the 3D printed micropillar shown in the SEM images in Fig. 3c aid in this process. Molecular structure of a rGO sheet is shown where -COOH and -OH groups are indicated. (c) Coupling of the viral antigens with the rGO sheets using EDC:NHS chemistry (step 3). The EDC and NHS molecules activate the -COOH group of rGO sheets. Recombinant antigens of the SARS-CoV-2 virus are immobilized and bound to rGO sheets by establishing strong covalent bonds between the -COOH group of rGO and -NH_2_ group of antigens via an amidation reaction. Two antigens, namely, spike S1 and RBD, were separately immobilized in this manner. A bovine serum albumin (BSA) treatment on the pillar surface blocked the non-specific sites of the sensor. (d) Antibodies selectively attached to the specific antigens upon introduction to the sensor via an antibody-antigen interaction (step 4). (e, f) Schematics showing the sensing principle of the 3DcC device. The electrode/electrolyte interface of the WE was expected to form an electrical double layer (C), inner Helmholtz plane or IHP, outer Helmholtz plane or OHP, and a diffusion layer during the redox reaction. An equivalent electrical circuit is shown. When antibodies are introduced (f), they rapidly bind with the antigens on the electrode surface, altering the Nyquist plot (schematics in e and f) which is captured by electrical impedance spectroscopy (EIS).

The 3DcC device works on the principle of electrochemical transduction.^26^ When an AC potential is applied between WE and RE, an electrical double layer is formed at the WE-electrolyte interface (Fig. 2e). For an electrolyte such as ferro/ferricyanide, the double layer can be formed due to accumulation of oppositely charged ions at the interface compared to the charge on the electrode (Fig. 2e). This creates a characteristic impedance (i.e., charge transfer resistance) when a constant potential is applied between, WE and RE. An equivalent circuit is shown in Fig. 2e, where C_dl_ is the double layer capacitance, R_ct_ is the charge transfer resistance, and R_s_ is the electrolyte resistance. Note that in the current study, the electrolyte used was phosphate buffer solution mixed with ferro/ferricyanide. When a fluid containing antibodies to the antigens shown in Fig. 2c are introduced in the microfluidic chamber, we expect selective binding of the target antibodies to the corresponding antigens on the electrode, which would increase the thickness of C_dl_ layer, causing a higher R_ct_, which could then be detected via electrochemical impedance spectroscopic (EIS) measurements (Fig. 2f). First, we expect the 3D geometry to accelerate the formation of the electrical double layer compared to a corresponding 2D surface, enabling a fast detection of the changes in R_ct_ and an increase in the sensitivity. Further, we expect that binding of the antibodies with antigens will increase the thickness of the double layer, causing the R_ct_ to increase proportionally. Thus, simple measurements of R_ct_ values would provide an excellent measure of the presence of antibodies in the fluid introduced in the 3DcC device.

### Physical Characterization of 3DcC Device

Figure 3 shows the microscopic and spectroscopic analysis of 3DcC device constructed using the process shown in Figs. 1 and 2. An optical micrograph of the device with CE, WE, and RE is shown in Fig. 3a. The scanning electron microscope (SEM) images of three-dimensional gold micropillar array electrode (i.e., WE) fabricated by AJ printing prior to rGO and antigen functionalization is shown in Figs. 3b and 3c. The diameter and pillar-to-pillar gap of the micropillars measured over 5 randomly chosen pillars per device across three devices was 73 μm (± 2.13%) and 122 μm (± 0.25%), respectively, while the height was about 250 μm. The top surface of solid pillar had a 15 μm deep and 20 μm diameter dip which was a result of the printing process depicted in the schematic of Fig. 1d. The surface texture of the gold pillars formed by nanoparticle sintering is shown in the zoomed-in SEM images of Fig. 3c and Fig. S1a. This texture consisted of micron-sized gold crystals on the pillar surface and a surface porosity caused by the nanoparticle sintering process. This surface texture was expected provide additional sites for rGO adhesion in addition to increasing the total surface area. The micropillar electrode after the rGO and antigen treatment (Fig. 2c) is shown in Figs. 3d, S1b, and S1c. Fig. 3d shows the rGO flakes attached to the gold micropillars as well as the gold base layer (also see Fig. S1c). The rGO sheets are attached to Au pillar surface non-uniformly due to their electrostatic interactions. Graphene flakes appeared wrinkled on the Au surface (zoomed-in images of Figs. S1b and S1c) which was expected to enhance the loading capacity of the antigens.

Raman spectroscopic measurements were conducted to investigate the defect (D) and graphitic (D) bands present in the rGO-Au micropillars (Fig. 3e) with and without antigens. The Au micropillar surface prior to rGO treatment did not show any Raman peaks. The Raman spectrum of rGO-Au micropillars revealed a D-band at 1348.1 cm^-1^ and a G-band at 1590.5 cm^-1^. The D-band (symmetry A1g mode) is due to the vibration of -sp^3^ carbon atoms or defects while G-band originates due to first-order scattering of E2g phonon of sp^2^ carbon atoms at the center of Brillion zone.^27^ The appearing D and G peaks in the Raman spectrum indicate the presence of rGO sheets on gold pillars. The intensity ratio (I_D_/I_G_) of rGO-Au was 0.84, which was slightly changed to 0.85 after incorporation of antigens. Antigen immobilization on rGO sheets influenced the peak position, which was shifted to higher wavenumber (5.5 cm^-1^) due to the decreased graphitic nature of the rGO sheets. Energy dispersive X-ray (EDX) studies indicated presence of large amount of carbon when rGO was coated on the gold micropillars (Fig. 3f and spectrum in Fig. S2). The existence of nitrogen for antigen immobilized sample (Fig. 3f and spectra in S2c) may come from the -NH**2** group of the protein (i.e. antigen). We note that the user interface for reading of the electrochemical cell signal was a laptop or a smartphone. A 3DcC device with a readout using a cellphone-based interface is shown in Fig. 3g.

**Figure 3.**
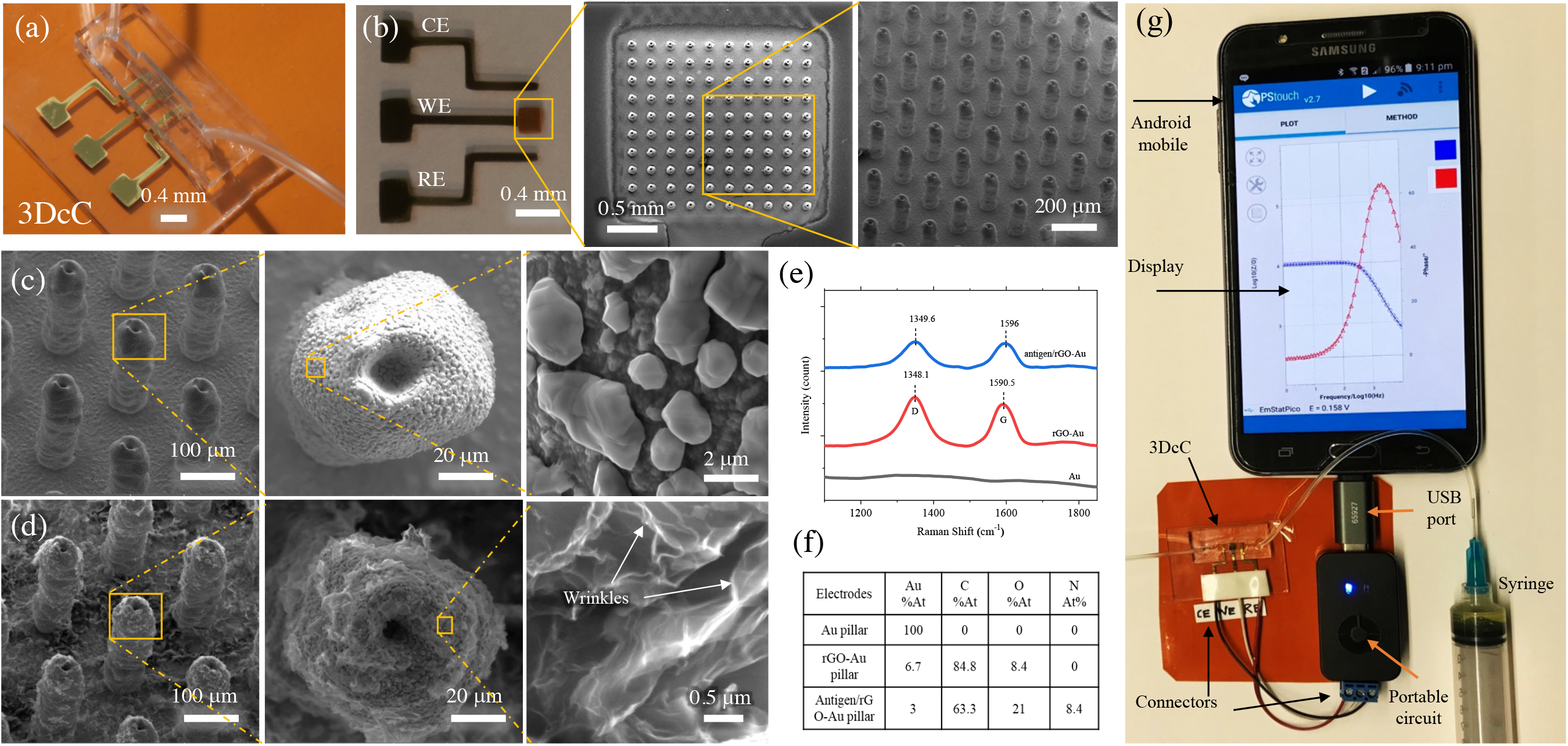
Physical and Chemical Characterization of the 3DcC device. (a) Optical image of the 3DcC device made by the fabrication process described in Figs. 1 and 2. (b) Optical and SEM images of the AJ printed gold micropillar array (10×10) at different magnifications. (c) SEM images showing the morphology of a single gold micropillar at different magnifications. The tips of the micropillars are pointed and have a small circular dip at the top (diameter 20 mm) which is a result of the printing process described in Fig. 1d. Surface morphology of the micropillars consisted of gold clusters that created a characteristic surface porosity. This morphology helps attach rGO flakes to the electrode surface. The diameter and pillar-to-pillar gap of the micropillars was 73 μm (± 2.13%) and 122 μm (± 0.25%), respectively, while the height was about 250 μm. (d) SEM images showing the morphology of a gold micropillar after rGO decoration. The rGO sheets have formed secondary three-dimensional networks adjacent to the micropillars. The rGO has also formed wrinkles due to n-n interactions between graphene sheets. The 3D micropillar geometry and the rGO coating are responsible for the increase in the loading of the viral antigens. (e) Raman spectra of an AJ printed gold micropillars without coating, after coating with rGO, and after immobilization of antigens on rGO-Au. The spectra show both defect and graphitic peaks for the coated samples, but no Raman peaks for the bare gold micropillar. The graphitic peak is shifted to higher number upon S1 antigen immobilization. (f) Energy-dispersive X-ray spectroscopy (EDX) studies showing atomic % before coating, after coating with rGO, and after immobilization of antigens on rGO-Au. (g) The 3DcC device interfaced with a portable Potentiostat (Palmsens Inc) which was connected to a smartphone via a mini-USB connection with the signal being read by PStouch software.

### Electrochemical Characterization of 3DcC Device

The 3DcC device shown in Fig. 3 was characterized by using cyclic voltammetry (CV) and electrochemical impedance spectroscopy (EIS). For comparison with the 3DcC device, we created a sensor with planar 2D WE, where the spike S1 antigen and bovine serum albumin (BSA) molecules were immobilized on a 2 mm × 2 mm planar Au surface (i.e., surface without AJ printed micropillars). For the 2D WE, however, we did not functionalize rGO on the surface as the rGO could not be bonded to the relatively smooth 2D gold surface which lacked surface porosity. Other experimental conditions for both sensors were kept identical. The 2D sensor showed low oxidation and reduction currents of a redox couple (ferro/ferricyanide) as shown in Fig. S3a of Supporting Information, which we believe was due to the immobilized spike S1 antibody and BSA molecules that blocked the electron transfer to the electrode from the electrolyte. Fig. S3a also shows that the 3DcC sensor (with rGO and S1 antigen coated gold micropillars) exhibits clear oxidation and reduction peaks of redox mediator with a 50× increase in current compared to the 2D electrode. Based on the Randles-Sevcik equation,^26, 28^ this significant increase in current cannot be explained by surface area alone (e.g., from pure geometry, the area of 3D electrode is 2.6× that of the 2D electrode). We speculate that since the two electrodes differ not only in geometry but also in surface chemistry, the diffusion of the electroactive species occurred differently for the two cases. For 3D electrodes, the geometry, in addition to an increase in the area, allowed radial or spherical diffusion, in addition to planar diffusion; compared to just planar diffusion for the 2D electrode. This was confirmed by COMSOL simulations described below. The effective diffusion coefficients for 3D electrodes will thus differ significantly when compared to the 2D electrodes.

The scan rate studies for sensors with 2D and 3D electrodes were carried out to assess the mass transport of potassium ferro/ferricyanide as shown in Figs. S3b and S3c of the Supporting Information. By varying the scan rate from 0.5 to 2 V/s, both the sensors were characterized (Fig. S3d). The anodic and cathodic peak currents for both sensors were directly proportional to their scan rates, but the voltages are shifted to more positive and negative potentials, respectively. This indicated that both the sensors exhibited a diffusion-controlled redox process.^29^ The Nyquist plots and phase shift (9) of sensors with 3D electrodes immobilized with recombinant SARS-CoV-2 spike S1 and RBD antigens and 2D electrodes immobilized with recombinant SARS-CoV-2 spike S1 antigens are shown in Fig. S3e and S3f, respectively. This data shows that the 3D electrodes exhibited a charge-transfer-limited process, whereas the 2D electrodes showed diffusion limited process (Warburg impedance) indicating a poor charge transfer characteristic.

In order to understand this phenomenon further, we carried out simulations of the CV experiments using COMSOL Multiphysics Software for the 2D and 3D electrodes (Fig. S4 of the Supporting Information). The bulk concentration was assumed to be 1 mol/m^3^, while the diffusion coefficient was taken from that obtained in the experiments from the Randles-Sevcik equation.^26^ Fig. S4a shows the electrode geometry, while Fig. S4b shows the diffusion profiles for a 10×10 array configuration. Fig. S4c shows 2D and 3D electrodes with their planar, and spherical and planar diffusion profiles, respectively. The concentration of the diffusing species for array configurations of planar and 10×10 3D electrodes are also shown in Fig. S4d. The presence of radial diffusion as shown in COMSOL simulations confirms the speculation that this phenomenon may help create high redox current in case of the 3D electrodes.

### 3D Sensing of COVID-19 Antibodies

SARS-CoV-2 S and N proteins have been used for serologic assays for detecting antibodies of SARS-CoV-2 infection.^14, 15^ The S protein binds to viral entry receptor angiotensin-converting enzyme-2 (ACE2) and mediates viral entry. It is present as a trimer with three receptor-binding S1 heads sitting on top of a trimeric membrane fusion S2 stalk.^30^ The receptor-binding subunit S1 contains the N-terminal domain (NTD) and the receptor-binding domain (RBD) while the membrane fusion subunit S2 contains the fusion peptide (FP), two heptad repeats (HR1 and 2), a transmembrane anchor (TM) and the intracellular tail (IC).^30^ Serologic assays using both recombinant S1 and RBD proteins have been shown to detect specific antibodies in COVID-19 patients.^31^ For antibody tests in this work, we thus chose the spike S1 and RBD antigens of SARS-CoV-2 to develop the 3DcC testing platform.^32^

Figure 4 shows the impedimetric sensing plots and charge transfer resistance (R_ct_) for the 3DcC sensor when the electrodes are exposed to phosphate buffer saline (pbs), rabbit serum (rs), fetal bovine serum (fbs), and spike S1 antibodies (rabbit IgG). The spike S1 antibodies were introduced successively at concentrations of 0.01 fM, 1 fM, 1 pM, 100 pM, 1nM, and 30 nM, three times each, wherein the sensor was regenerated after each set of measurements, including the controls testing. In the first set of measurements (Fig. 4a), a testing baseline was set by allowing only the pbs solution with no target antibodies which showed a charge transferred-limited process with R_ct_ of 3.51 kW. While the sensor was tested with biological samples such as rs and fbs as controls, the sensor generated similar profiles of Nyquist plots having a deviation of ± 6.01% compared to the baseline signal. The adsorption of any proteins or albumins can change the sensor signals significantly. However, though these proteins have a major composition of albumin protein, the sensor did not show a significant change in the baseline signal. The sensor surface contained a layer of BSA molecules which are negatively charged, which may have repelled the albumin molecules in the serum due to their identical charge polarity.^29^ This result indicated that the sensor was selective to specific proteins as was desired. With these control measurements, the 3DcC device was tested with low concentrations of spike S1 antibodies at 0.01 fM and 1 fM. With the 0.01 fM concentration, a change of impedance signal (4.2 kΩ) was observed (Fig. 4d) compared to the sensor baseline and the control serum. This is as expected since binding of the antibodies with the antigens will obstruct electron transfer from the electrolyte to the electrode surface resulting in an increase in circuit impedance as captured by the Nyquist plot. Between 0.01 fM and 1 fM, the sensor did not show any significant change in impedance signal. Within this range of concentration of antibodies, the sensor surface may have only limited number of antibodies resulting in an unchanged signal. At 1 pM concentration of antibodies, however, the sensor showed a significant increase in the impedance to 5.21 kΩ. As the antibody concentration was increased to 1 nM, the R_ct_ increased further as reflected in the Nyquist plots. Interestingly, when a 10 nM concentration was tested, the sensor provided a very significant increase in the impedance signal (12.3 kΩ). Further, a minute change of signal (12.7 kW) was observed at 30 nM concentration. Testing beyond 10 nM concentration of spike S1 antibodies showed that the sensor exhibited a saturated impedance signal, wherein the maximum number of binding sites on the sensor surface are likely to be occupied by the target antibodies.

**Figure 4.**
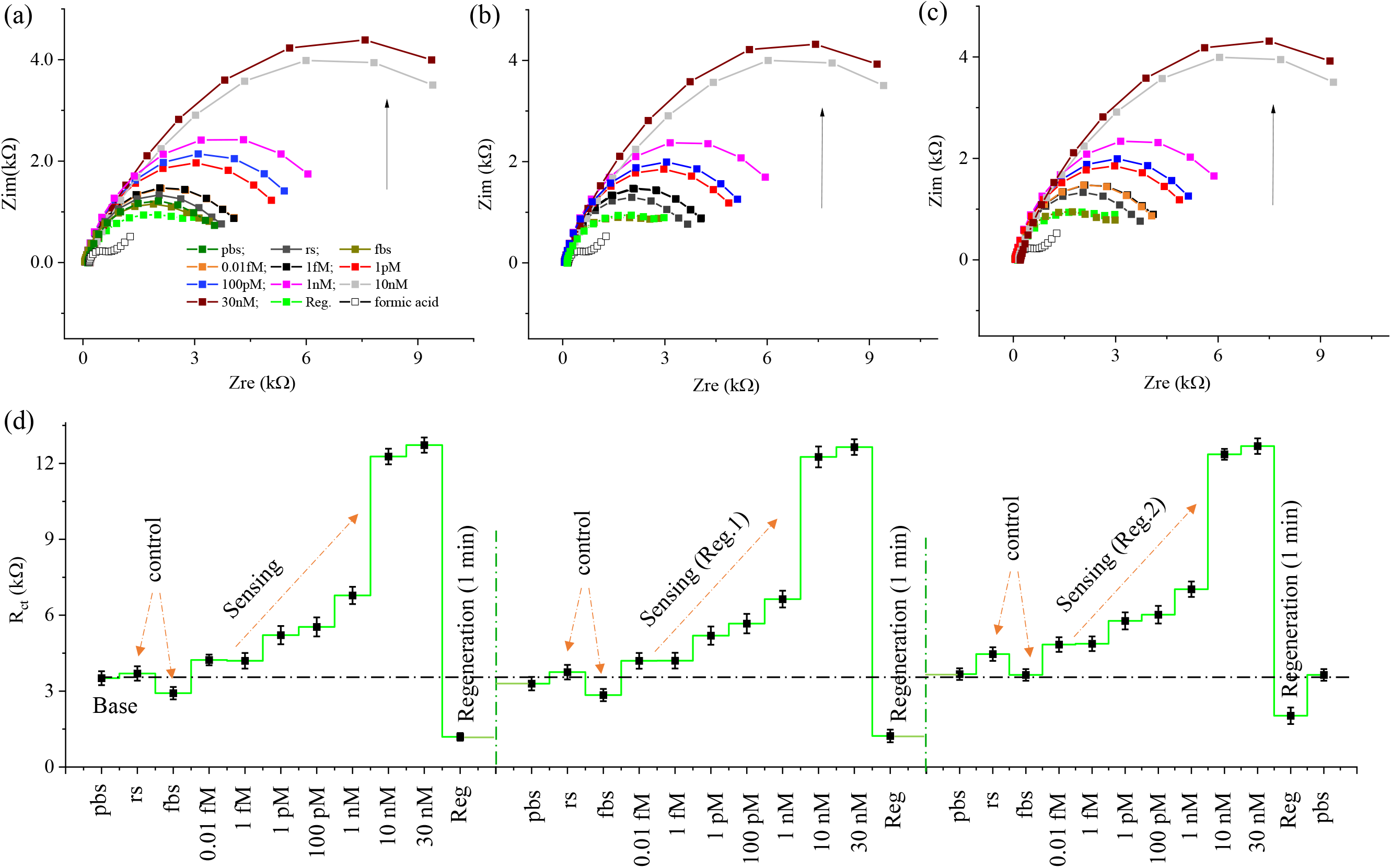
Sensing of Antibodies to SARS-CoV-2 Spike S1 Antigen at Different Molar Concentrations with Regeneration. (a) Nyquist plots of the 3DcC sensor measured by EIS method without and with the spike S1 antibodies at concentrations of 0.01 fM, 1 fM, 1 pM, 100 pM, 1 nM, 10 nM, and 30 nM in PBS buffer solution. (b, c) Nyquist plots similar to that in (a) after two successive sensor regenerations by a low pH chemistry consisting of 1.0 M (pH 2.5) formic acid solution. The regeneration was achieved within 60 seconds. For all concentrations, the signal in (b, c) was within 95% of that in (a). (d) The charge transfer resistance (R_ct_) for the 3DcC sensor for each concentration of antibodies and control serum before and after each regeneration for the data in (a, b, c). The fetal bovine serum (fbs) and rabbit serum (rs) in (a-d) were utilized as control biofluids. For all measurements, a 50 mM PBS (pH 7.4) solution containing an equimolar concentration (5 mM) of a ferro/ferricyanide mediator was used. Three successive readings were obtained at each concentration of the antibodies for the three data-sets. Detection frequencies from 1 Hz to 10,000 Hz were applied to obtain this data. There was no incubation time for all the measurements in this figure. The R_ct_ values in (d) were calculated by fitting the data in (a, b, c) to a Randles equivalent circuit shown in Fig. 2e.

To elute antibodies from the sensor surface, we exposed the sensor electrodes to a solution of formic acid (1.0 M) having a pH of 2.5. This was chosen as a pH of 2.5 is outside the physiological range for human body (7-7.4 pH), where antibodies are expected to elute from the antigens via a disruption of immunoaffinity.^33^ Indeed, upon introduction of the formic acid solution for 60 seconds into the sensor, the R_ct_ values dropping to about 1.2 kW. In addition to eluting the antibodies, the low R_ct_ is caused by high electron transfer rates in acidic media. Note that the signal from formic acid in Figs. 4a, 4b, and 4c are after the 1^st^, 2^nd^, and 3^rd^ regeneration. After the 60 second incubation by formic acid solution, the sensor was washed with pbs solution. At this point, the sensor recovered to 94% of its base signal. Figs. 4b and 4c show sets of measurements after regeneration for the same set of spike S1 antibody concentrations as studied in Fig. 4a. After 1^st^ regeneration, the signal with rb and fbs (Fig. 4b) was within ±6.96% of the baseline. After 2^nd^ regeneration, this signal did not change significantly (Fig. 4c). We also observed similar patterns of impedance signals for spike S1 antibody solutions at different concentrations after 2^nd^ regeneration as shown in Figs. 4c and 4d. The sensor could provide a low LoD of 1 pM for detection of spike S1 antibodies.

Figure 5 shows the sensing of RBD antibodies using the 3DcC device where results are presented in a similar manner as that for spike S1 antibodies (Fig. 4). The device had micropillar electrodes with immobilized SARS-CoV-2 RBD-His recombinant antigens. The EIS spectra for each concentration of RBD antibodies with control biofluids such as rs and fbs are shown in Fig. 5a; while that after two successive regenerations are shown in Figs. 5b and 5c. The RBD antibody concentrations were set from 1 fM to 10 nM to test the sensor. The sensor base signal was obtained by collecting impedance spectra in presence of pbs solution where the sensor showed a R_ct_ of 3.89 kW (Fig. 5d). With the rs and fbs, the sensor showed a deviation of 3.0% in the impedance after regeneration when compared to sensor baseline. The sensor did not show any increased signal when 1.0 fM concentration of RBD antibodies were introduced. At RBD antibody concentration of 1 pM, the R_ct_ value changed to 5.07 kW. Similar to the spike S1 antibodies, as the concentration of RBD antibodies was increased from 1pM to 10 nM, the R_ct_ values were found to increase as shown in Fig. 5d. The regeneration of the RBD sensor was achieved by 1.0 M formic acid (pH 2.5) solution in 60 seconds; the same as that used for the regeneration of spike S1 antibodies. Unlike the spike S1 sensor, the RBD sensor showed different linear responses between 1 pM to 100 pM, and 100 pM to 10 nM. After two regenerations, a minimum loss of sensitivity (± 1%) was observed for the RBD sensor. Based on the results in Figs. 4 and 5, we created a detection band for spike S1 and RBD sensing, respectively, where a minimum R_ct_ value was identified above which the sensing could be achieved (see Fig. S5a and S5b of Supporting Information).

**Figure 5.**
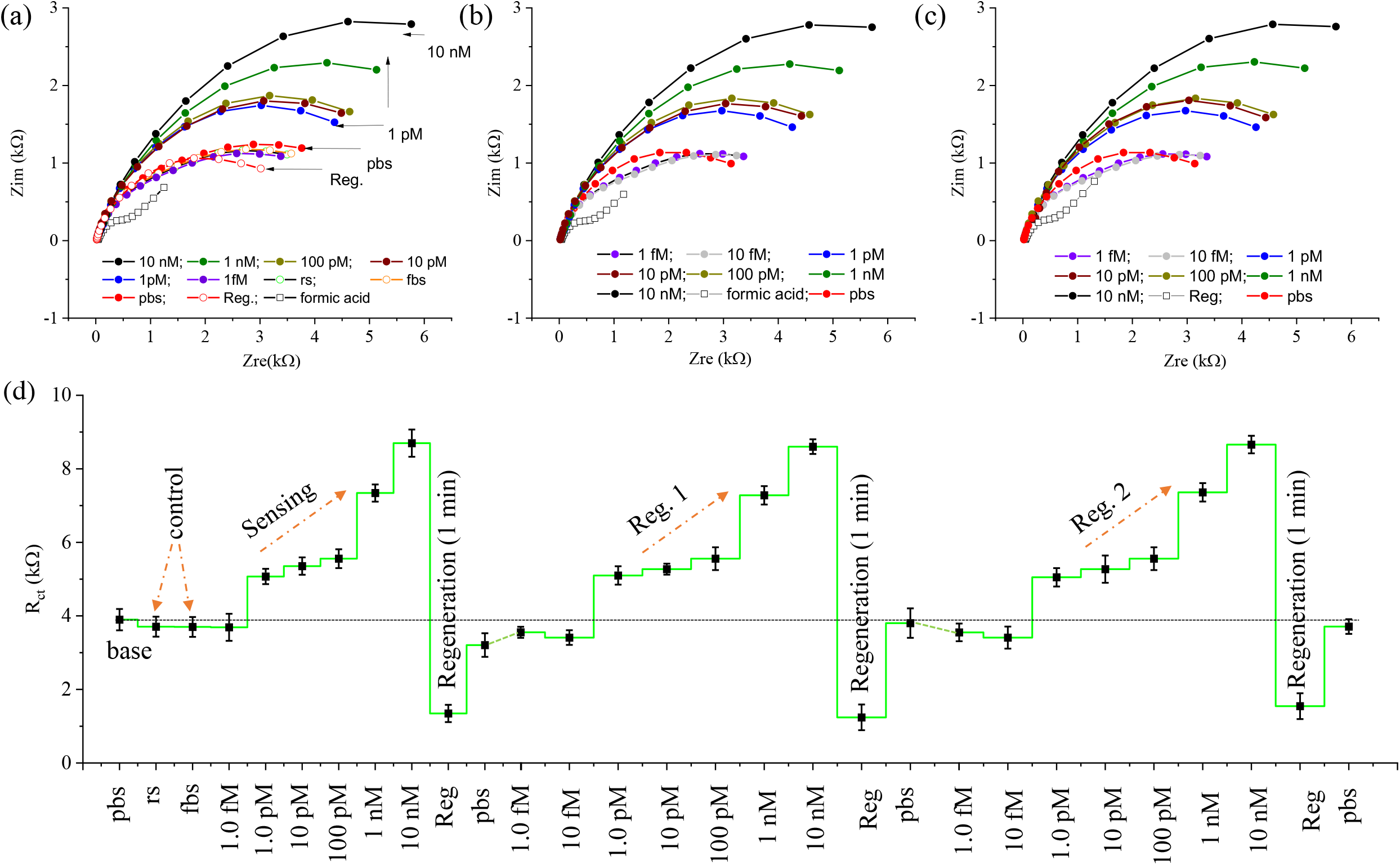
Sensing of Antibodies to SARS-CoV-2 receptor binding domain (RBD) Antigens at Different Molar Concentrations with Regeneration. (a) Nyquist plots of the 3DcC sensor measured by EIS method without and with the RBD antibodies at concentrations of 0.01 fM, 1 fM, 1 pM, 100 pM, 1 nM, 10 nM, and 30 nM in PBS buffer solution. (b, c) Nyquist plots similar to that in (a) after two successive sensor regenerations by a low pH chemistry consisting of 1.0 M (pH 2.5) formic acid solution. The regeneration was achieved within 60 seconds. For all concentrations, the signal in (b, c) was within 95% of that in (a). (d) The charge transfer resistance (R_ct_) for the 3DcC sensor for each concentration of antibodies and control serum before and after each regeneration for the data in (a, b, c). The fetal bovine serum (fbs) and rabbit serum (rs) in (a-d) were utilized as control biofluids. For all measurements, a 50 mM PBS (pH 7.4) solution containing an equimolar concentration (5 mM) of a ferro/ferricyanide mediator was used. Three successive readings were obtained at each concentration of the antibodies for the three data-sets. Detection frequencies from 1 Hz to 10,000 Hz were applied to obtain this data. There was no incubation time for all the measurements in this figure. The R_ct_ values in (d) were calculated by fitting the data in (a, b, c) to a Randles equivalent circuit shown in Fig. 2e.

We next discuss the detection time of the 3DcC sensor for spike S1 antibodies and RBD antibodies. Fig. 6a show the impedance of the sensor as a function of time in seconds for the detection of spike S1 antibodies. It is noticed that with no target in the pbs solution, the sensor impedance started to change at 3 seconds and reached to ~1 kW in 10 seconds, beyond which it became saturated. When spike S1 antibodies (1 nM) were introduced in six different sensors, a significant change of impedance was observed (in line with data in Fig. 4) and reached about 93.2% of the saturation value in 11.5 seconds, indicating detection of antibodies. The magnitude of impedance at 11.5 seconds as a function of the spike S1 antibody concentrations (0.01 fM to 30 nM) for the 3DcC device is shown in Fig. S6a of Supporting Information. The Fig. 6b shows the impedance as a function of time for six different 3DcC RBD sensors when RBD antibodies are introduced at 1 nM concentration. For all the sensors, the signal reached 92% of the saturation signal at 11.5 seconds, again indicating detection of the antibodies. Fig. S6b shows the time required to reach 95% of the saturation signal for the data presented in Figs. 6a and 6b. *The 3DcC device can thus detect the antibodies to SARS-CoV-2 virus within seconds, which is faster than any data yet reported in literature*.

**Figure 6.**
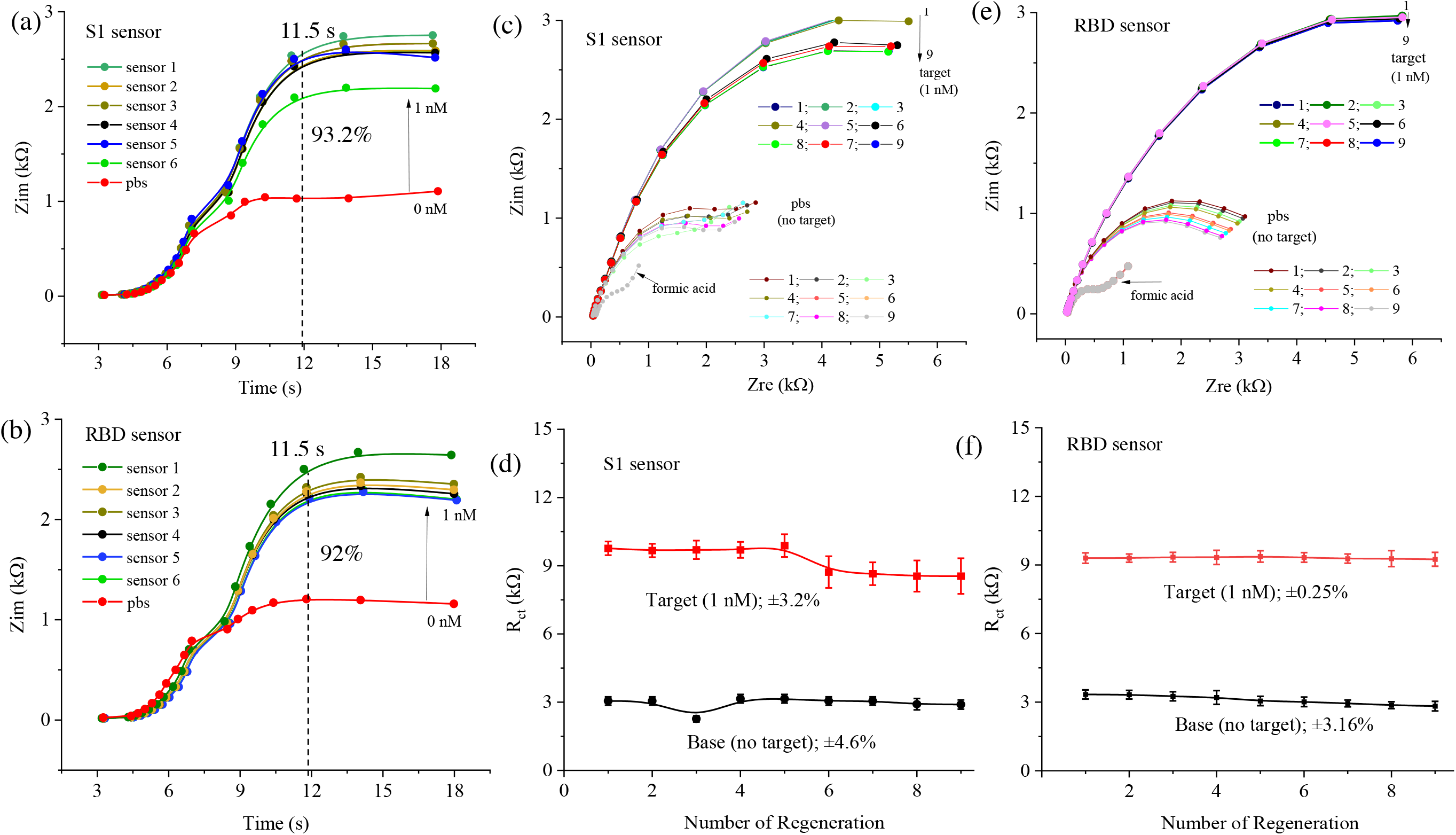
Detection Time and Regeneration Studies of the 3DcC Device. (a, b) The detection time for spike S1 and RBD antibodies during the EIS measurements, respectively, for twelve different 3DcC devices. The device impedance is plotted against the detection time. The spike S1 and RBD sensors reached 93.2% and 92% of their saturation impedance at 11.5 seconds, respectively, allowing the signal detection in seconds. The concentration of antibodies was set to 1 nM for this measurement. A frequency range of 1-10000 Hz was used to obtain this data. The (Zim) data is recorded from 3 seconds after the introduction of antibodies as the EIS measurement had to overcome the solution resistance, R_s_ prior to obtaining the charge transfer resistance (see schematic in Fig. 2e). (c, d) Regeneration study showing Nyquist plots and charge transfer resistance for the detection of spike S1 antibodies for the 3DcC device. The regeneration is carried out 9-times (a total of 10 readings). For each measurement, the sensor was first exposed to pbs, then to 1 nM concentration of S1 antibodies in pbs, and finally to formic acid (1M) with an incubation time of 60 s. The charge transfer resistance as a function of the number of regenerations is plotted in (d). (e, f) Regeneration data for sensing of RBD antibodies by the 3DcC device. For both the sensors, a minimal loss in sensor performance is observed after 9 regenerations.

To validate the sensor regeneration phenomenon, 3DcC sensors for the detection of spike S1 antibody and RBD antibody were regenerated 9× each as shown in Figs. 6c and 6d, and Figs. 6e and 6f, respectively. A pbs solution was used as a reference while 1 nM antibody solutions were used as the target. After 9× regenerations, the spike S1 and RBD sensors did not show significant changes in the R_ct_ values as evidenced by their low relative standard deviation (RSD) of less than ±3.2 *%* and ±0.25 %, respectively. However, the base line R_ct_ of spike S1 and RBD sensors was found to change by ±4.6% and ±3.1% when compared to that with no regeneration. We noted that even after 9 regenerations (i.e., 10^th^ reading), both sensors provided a good signal, indicating a high regeneration capability. We speculate that acidic media may degrade the bonding between rGO and antigens after repeat exposures, possibly limiting the regeneration capability beyond a certain limit.

The sensitivities of the 3DcC device for spike S1 and RBD sensors are shown in Figs. S6c and S6d of the Supporting Information. These plots are obtained from the R_ct_ values in Figs. 4d and 5d. For spike S1 sensor, the slope of Fig. S6c was 0.27 ±0.04 kΩ/nM in the range of 1.0 fM to 1 nM, and 4.5 ±1.1 kΩ/nM in the range of 1 nM to 30 nM. The high sensitivity at higher concentrations is likely due to higher number of antibodies captured by the sensing surface. For RBD sensors the slope of Fig. S6d was 3.9 ±0.04 kΩ/nM for a range of 1.0 fM to 0.1 nM, and 1.6 ± 0.16 kΩ/nM for a range of 0.1 nM to 10 nM. The spike S1 sensor showed a higher sensitivity at a higher concentration of antibodies when compared to RBD sensor. We speculate that this high sensitivity of the 3DcC device was due to the 3D architecture, a high porosity, and specific surface chemistry that allowed an enhanced loading capacity of the antigens.

Figs. 7a and 7b show the Nyquist plots and R_ct_ for cross-reactivity study of the 3DcC sensor designed to test spike S1 antibodies, respectively. The sensor was tested in the presence of RBD antibodies, nucleocapsid (N) antibodies, and cytokines such as interleukin (IL)-6 antigens, without and with the spike S1 antibodies (1 nM). In the absence of spike S1 antibodies, the sensor was found to be insensitive to N antibodies, RBD antibodies, and IL-6 protein molecules as evidenced by its low R_ct_ values and low RSD (±6.9%). The sensor showed a slight deviation (±6.5%) from the initial signal in the presence of RBD antibodies suggesting the sensor was either saturated by the S1 antibodies. Alternatively, the RBD antibodies might recognize linear epitope while the S1 protein might have a specific confirmation that prevents binding to the RBD antibodies. The sensor was highly sensitive when the spike S1 antibodies were introduced with the above molecules, leading to an increase in the R_ct_ values s shown in Fig. 7b. Figs. 7c and 7d show the cross-reactivity studies of RBD antibodies. Again, we used N antibodies, IL-6 antigens, and spike S1 antibodies in the absence and presence of RBD antigens (1 nM). In this result, the 3DcC device tested with IL-6 antigen showed a deviation of ±11.0 % compared to base line with no target. We note that the decrease of R_ct_ value with IL-6 antigen did not affect the sensor response when target antibodies were present in the buffer solution. Without RBD antibodies, the 3D sensor showed an RSD of ± 5.81 % compared to baseline signal while the RSD was reduced to ± 2 % in the presence of 1 nM target RBD antibodies. It is clear that specific binding sites on the antigen molecules (epitope) reject binding by non-specific molecules but allow binding to a specific antibody (paratope). These results indicated that the sensor showed a good selectivity even within a similar group of proteins.

We next show the sensor reproducibility (Figs. 7e-7h) where twelve 3DcC sensors, 6 for spike S1 antibodies and 6 for RDB antibodies were evaluated. Nyquist plots were obtained and R_ct_ values were calculated at fixed concentration (1nM) of spike S1 (Figs. 7e and 7f) and RBD (Figs. 7g and 7h) antibodies. For spike S1 and RBD antibodies, the sensors showed a RSD of ± 2.7% and ± 2.5%, respectively, indicating reasonable reproducibility of the sensor. Note that additive manufacturing (AM) processes are digital (i.e. controlled by CAD programs) and hence are expected to be repeatable. However, all AM processes are known to have microstructural inhomogeneities in their final parts.^34^ It is clear that for the 3DcC device, the AJ nanoparticle printing process created repeatable structures (Fig. 3b, S1a) that provided acceptable repeatability in antibody sensing performance (Figs. 7e-7h), but led to a microstructure on the pillar surface (high magnification images of Figs. 3c and S1a) that aided in the surface functionalization process.

**Figure 7.**
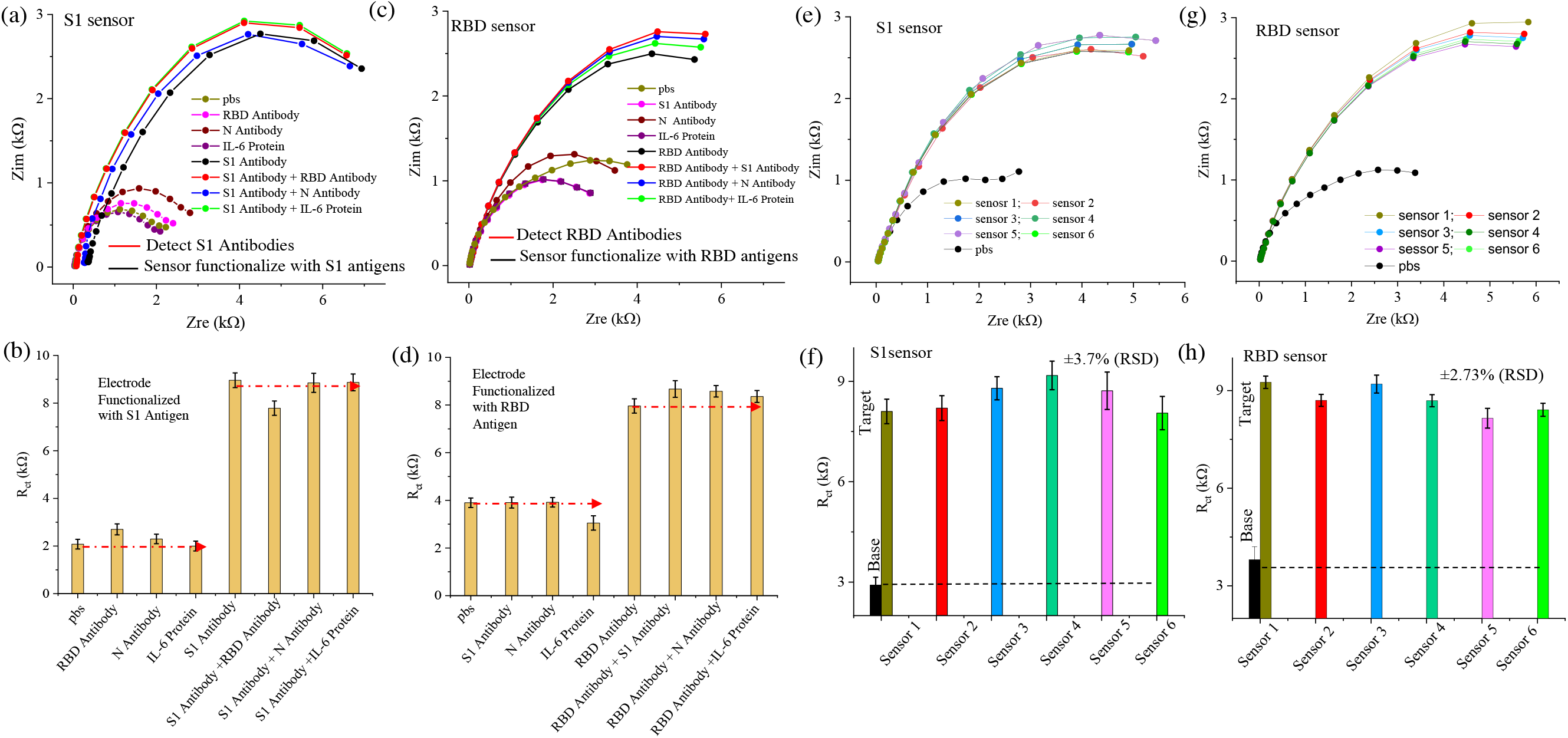
Cross-reactivity and Reproducibility Studies of the 3DcC Device. (a) Cross-reactivity test for the 3DcC device designed to detect spike S1 antibodies. Nyquist plots for the device are plotted for multiple antigens and antibodies in absence and presence of spike S1 antibodies. Interleukin-6 or IL-6 antigen (0.05 nM), nucleocapsid (N) antibody (1 nM), and RBD antibody (0.1 nM) are used for this measurement. (b) The R_ct_ values were calculated from the Nyquist plots shown in (a). The 3DcC device provided minimal interference with other related proteins. (c) Cross-reactivity test for the 3DcC device designed to detect RBD antibodies. Nyquist plots for the device are plotted for multiple antigens and antibodies in absence and presence of RBD antibodies. Interleukin-6 or IL-6 antigen (0.05 nM), nucleocapsid (N) antibody (1 nM), and spike S1 antibody (0.1 nM) are used for this measurement. (b) The R_ct_ values were calculated from the Nyquist plots shown in (a). The 3DcC device provided minimal interference with other related proteins. (e, f) The sensor reproducibility test on six different 3DcC sensors in presence of S1 antibodies (1nM in pbs). The sensor-to-sensor variation is evaluated by calculating the R_ct_ values for each sensor. This variation is within about 6%. The error bar is from at least three repeated measurements of the sensor. (g, h) Reproducibility test data for sensing of RBD antibodies (1 nM in pbs) from six different sensors. This sensor-to-sensor variation in this case was about 5%.

We also investigated the real-time tracking of binding kinetics of antigens and antibodies at the sensor surface. Figs. 8a-8c show R_ct_ values, impedance graphs, and a schematic explaining the various phases of binding events such as association, dissociation, and regeneration, respectively. In the association phase, the sensor is exposed to a concentration of target antibodies (spike S1 in this case) where both bound and unbound molecules were present resulting in a slightly higher R_ct_ (Fig. 8a). In the dissociation phase, the sensor was washed by buffer solution and unbound antibodies were removed from the sensor resulting in a slight reduction in the R_ct_. However, during regeneration by a low-pH chemistry, elution or desorption of antibodies from the antigens occurred via disruption of ionic and hydrogen bonds between in the immunocomplex,^35, 36^ reducing the R_ct_ to a low value. Quantification of the minute changes in impedance behavior thus leads to a sensitive tracking of both association and dissociation phases of the 3DcC sensor operation.

**Figure 8.**
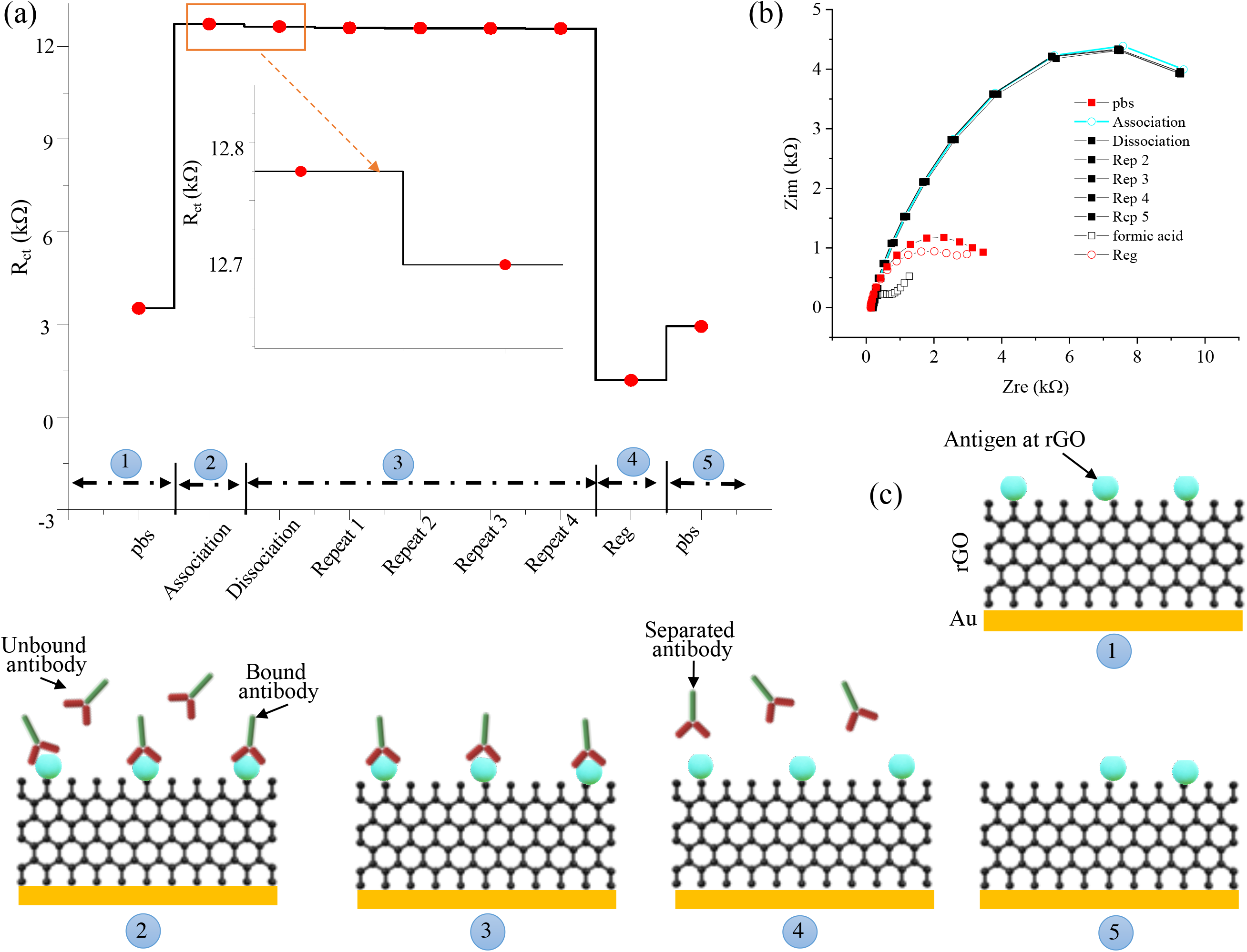
Study of the Kinetics of Antigen-Antibody Interactions. (a) Variation in the charge transfer resistance (R_ct_) during antigen-antibody interaction for spike S1 antibodies at the micropillar surface indicating their association, dissociation, and regeneration. (b) Nyquist plots for the data in (a). (c) Schematic of the association, dissociation, and regeneration of the kinetics shown in (a, b). In associate phase, the target antibodies (30 nM) with buffer solution are loaded into the 3DcC device. The antibodies attach to the antigens. However, some unbound antibodies may be present in the solution near the sensor surface and contribute to the R_ct_. In dissociation phase, a fresh pbs (without antibodies) is used to wash the 3DcC device, thus removing the unbound antibodies in the solution, and reducing the R_ct_ slightly. In the regeneration phase, a solution of formic acid (1 M; pH 2.5) is used to regenerate the sensor surface, thus eluting the antibodies from the antigens and lowering R_ct_ significantly. Five repeated measurements are carried out during the dissociation phase.

## Discussion

The AJ printed biosensing platform (i.e. 3DcC device) developed in this work can detect antibodies for SARS-CoV-2 within seconds. This result is highly significant in pandemic control and will positively impact public health. The research also represents an important advance in biotechnology where 3D printing is used to realize architected electrodes that enable rapid detection of pathogenic biomarkers at low concentrations.

The 3DcC device (Fig. 3), which includes the WE with micropillar geometry and surface porosity, and a chemistry involving flaky rGO (Fig. 3 and S1b) allowed a high redox current for the electrochemical cell forming the sensor (Fig. S3a). This increase was due not only to the larger surface area of the 3D electrode but also to the presence of radial and linear diffusion in the 3D microelectrode geometry (Fig. S4c). Further, the 3D geometry allowed a relatively high loading of biorecognition elements (antigens) for the sensor, enabling the detection of the antibodies at concentrations as low as 1 pM and 1 fM for spike S1 and RBD antibodies (Figs. 4 and 5), respectively. Figs. 7a-7d demonstrate that the natural construction of antigen-antibody immunocomplex makes sensing selective. Another important outcome of this work is the regeneration of the sensor in one minute. By introducing formic acid with a pH of 2.5, which is outside of the physiological range observed in the body (7-7.4), the antibodies were eluted from the antigens allowing full recovery of the original sensor signal. The demonstration of 10 repetitions (9× regenerations) of the sensors for each of the antibodies (Fig. 6c-6f) gives the opportunity for the same sensor to be used multiple times in the field without losing the specificity and sensitivity. We also note that the 3DcC requires only a few μL of fluid for detection of antibodies (e.g. volume obtained by a finger prick). Lastly, a convenient cell phone-based user interface (Fig. 3g) will enhance testing accessibility of the device in underdeveloped areas.

We point to several advantages offered by the 3D printing method used to manufacture the micropillar electrodes of the device. The AJ nanoparticle 3D printing is a digital manufacturing platform where the sensor design is controlled by CAD programs. The fabrication is also rapid – the 10×10 gold micropillar array (Fig. 2b) takes 35 minutes to print per printhead. With the availability of AJ printers with four printheads working in tandem, the effective printing time can be reduced to 7-8 minutes per sensor followed by a batch sintering process. Such digital manufacture allows fabrication in two simple steps without the need for any cleanroom processes, lowering resources/cost per sensor that can improve accessibility of testing. In addition, 3D printing enables the sensor design to be changed via simple changes to CAD programs. A multiplexed chip where several working electrodes are printed on the same device can allow selective immobilization of several antigens of SARS-CoV-2 to detect multiple antibodies at the same time. Such a device can improve fidelity of detection and also allows the same device to be used for multiple tests. Such work will be part of a future investigation. The microstructural features (Fig. 3c and S1a) of the gold micropillars are a result of the AJ printing process and allows an excellent coverage of the rGO on the electrode surface (e.g. rGO could not be coated on a smooth gold surface). In addition, rGO forms a secondary 3D structure (Fig. S1b), allowing high loading of the antigens. We also note that the EDC:NHS chemistry used in the work covalently binds the -COOH group of rGO to -NH**2** group of antigens, which is common to several proteins. It is thus feasible to immobilize antibodies on the sensor surface and detect pathogens directly via their antigens, which will be part of a future investigation

The 3DcC device has several advantages over other methods for COVID-19 detection. Genomic sequencing, CRISPR-based test, and RT-qPCR^8-10^ are accurate but have a long testing time (days). Other methods such as ELISA and lateral flow measurements are less sensitive and cannot provide diagnostic information within seconds at high accuracies.^14, 15^ The 3DcC device, however, has the ability to detect SARS-CoV-2 antibodies non-destructively within seconds at a low LoD (down to a pM concentration) with label-free probing of antibodies and an ability for regeneration within one minute. These results are highly encouraging and a device for field use, after appropriate human trials, will have a significant positive impact on public health and the course of the current^37^ as well as future pandemics. Note that we obtained a signal in all the 3DcC devices used in this research every time when an antibody solution was introduced (Figs. 4-8). Although this is a very good initial indicator, studies with large scale patient samples to evaluate the sensitivity and specificity are necessary in order to establish statistical rates for false negatives and false positives. We also note that the concentration of antibodies to SARS-CoV-2 in body fluids of COVID-19 patients likely varies significantly (e.g. for asymptotic vs symptomatic patients) and has not been investigated at this time. However, 1 femto or 1 picomolar limit-of-detections for antibodies may enable an early detection of the disease which needs to be investigated urgently.

In summary, we have developed a 3D printed biosensing platform to detect antibodies specific to SARS-CoV-2 within seconds. The device is fabricated by using Aerosol Jet nanoparticle 3D printing method to create gold micropillar array electrodes, followed by functionalization of rGO and immobilization of antigens on the electrode surface using an EDC:NHS chemistry. The detection of antibodies was achieved by electrochemical transduction when antibodies introduced in a fluid formed an immunocomplex with antigens on the 3D electrode surface. This signal was highly selective and repeatable. Antibodies to spike S1 and RBD antigens of the virus were detected at concentrations of 1 pM and 1 fM, respectively. A low pH chemistry was used to regenerate the sensor via elution of antibody-antigen immunoaffinity within 1 minute, allowing up to 10 successive readings from the same sensor with high fidelity. This biosensing platform will allow rapid detection and early isolation of infection, saving lives. The test platform is generic and can potentially be used to detect biomarkers for other pathogens. The research represents a significant advance in biotechnology and will have a positive impact on public health.

## Materials and Method

### Materials

To construct the three-dimensional array electrode using AJ printing, we used a commercial gold (Au) nanoparticle ink (UTDAu40, UT Dots Inc., Champaign, IL). The average Au particle size was 4 nm, the ink viscosity was 3 cP, and particle loading in the ink was 40 wt%. The Au nanoparticles were dispersed in an organic non-polar solvent, which was aerosolized during AJ printing via ultrasound energy. The PDMS (SYLGARD™ 184 Silicone Elastomer Kit, Dow Corning, Midland, MI, USA) mixed with a base to hardener ratio of 10:1 to create the microfluidic channel of the 3DcC device.

Human recombinant SARS-CoV-2 spike S1-His protein (50 p,g/mL) and SARS-CoV-2 spike receptor binding domain (RBD)-His protein (50 μg/mL) expressed in HEK293 cells, were the antigens purchased from the Sino Biological US Inc., Wayne, PA. Before immobilizing on the 3D electrode surface, both the antigens were diluted to 5 p,g/mL using the carbonate buffer solution (pH ~9.6). Two rabbit immunoglobin (IgG) antibodies, SARS-CoV-2 spike S1 antibody (10 μL), and SARS-CoV-2 spike RBD antibody (10 μL) were also obtained from Sino Biological US Inc., Wayne, PA. Both the antibodies were diluted in phosphate buffer saline solution (pH 7.4) containing a 5 mM ferro/ferricyanide before their introduction into the microfluidic channel for measurement. These solutions were stored in −20°C before their use. Mouse monoclonal antibody (MAb) of human recombinant SARS-CoV-2 nucleoprotein (Cat. No. 40143-MM05) was purchased from Sino Biological Inc., Wayne, PA. *E. coli* derived human recombinant interleukin-6 (IL-6; Cat. No., 206-IL) antigen was purchased from R&D Systems, Inc., Minneapolis, MN. Bovine serum albumin (BSA), sodium bicarbonate, sodium carbonate, formic acid, phosphate buffered saline powder, EDC (1-ethyl-3-(3-dimethylaminopropyl) carbodiimide hydrochloride) and NHS (N-hydroxysuccinimide) were acquired from Sigma Aldrich, St. Louis, MO. A room temperature curable silver/graphene conductive epoxy (type G6E-RTSG, Graphene Supermarket, Inc., Ronkonkoma, NY) was used to connect wires to the pads of WE, CE, and RE (Fig. 1) of the 3DcC device.

### Fabrication of Electrodes for 3DcC Device

Figure 1 shows the fabrication process of the 3DcC device for rapid detection of COVID-19 antibodies. First, a patterned Au layer (with Cr adhesion layer) was deposited on a glass slide (Fig. 1a) which formed the base for the three electrodes, namely, RE, WE, and CE. To pattern Au/Cr layer, a shadow mask was created using a Kapton tape which cut using an automated cutter (Silhouette Curio™, Silhouette America®, Inc., Lindon, UT) with the aid of AutoCAD software (AutoCAD 2015, Autodesk Inc., San Rafael, CA). A 5 nm thick Cr layer was deposited as an adhesive layer, followed by a 100 nm thick Au layer using an e-beam evaporator (Kurt Lesker PVD 75, Jefferson Hills, PA) while using the Kapton shadow mask. The area of the patterned WE was 2 mm x 2 mm. Aerosol Jet nanoparticle 3D printing was then used to fabricate the three-dimensional micropillar electrode on the WE as shown in Figs. 1b-d.

The schematic of the AJ 3D printer (Model AJ-300, Optomec, Inc., Albuquerque, NM USA) is shown in Fig. 1b. The AJ printer consisted of an ultrasound atomizer, a deposition head with a nozzle, a movable and platen (X-Y direction) with temperature control, and a shutter to break the flow of the aerosol as necessary. The ultrasonic atomizer created a mist of aerosol droplets of about 1-5 im diameter from the Au ink, with each droplet containing the 2-5 nm Au particles. A carrier gas (N**2**) transported the droplets to the nozzle of the deposition head, while a sheath gas (also N**2**) helped focus the nanoparticle beam to a length scale of 10 μm. The carrier gas pressure was set to be about 24 sccm and sheath gas pressure to set to 60 sccm. The nozzle diameter used for printing was 150 μm. Before printing, the geometry of the micropillar array was drawn in AutoCAD using a program in the software AutoLISP (AutoCAD 2015, Autodesk Inc., San Rafael, CA) and converted to a “prg” file compatible with the AJ printer software. One milliliter gold nanoparticle ink without any dilution was loaded inside a glass vial (Fig. 1b), atomized using ultrasonic energy, and transported to the AJ printer. An external heating element was placed on top of the X-Y stage to heat the substrate to 150 °C. A layer-by-layer printing sequence was used to build up the 3D 10 x10 micropillar arrays from AJ droplets where the path of the droplets could be periodically blocked (or cleared) by controlling the shutter. The schematic of complete micropillar array is shown in Fig. 1c. The printing of an individual pillar is shown in Fig. 1d, where a layer of droplets was deposited on top of the previously printed and solidified Au micropillars. The strong surface tension of the printed ink caused the pillar to form without any support structures (Fig. 1d). The printing process for the 10×10 micropillar array was completed within 35 min for a given printhead. After printing, the dried micropillars array was heated to 400 °C for 5 h to completely remove the binders and sinter the Au nanoparticles which completed the manufacture of the WE of the device.

The surface of the RE on the glass substrate (Fig. 1a) was then coated with commercial silver/silver chloride ink (Ercon, Inc. Wareham, MA) by using a Kapton shadow mask. The curing temperature for this coat was 150 °C for 2 hr. The ink composition comprised of dispersed chloridised silver flake particulates in a solvent. Compared to thin film Ag/AgCl reference electrode, the commercial ink was chosen due to its improved stability as demonstrated in literature.^38^

The process of functionalization of WE are shown in Fig. 2. First, the gold micropillars were functionalized with rGO (Fig. 2a and 2b). The rGO sheets were obtained in powder form (CAS-No. 7782-42-5, ACS Materials LLC, Pasadena, CA). The rGO sheets were dispersed in DI water (0.2 mg/mL) and sonicated for 2 h. As per the manufacturer, the rGO was obtained from graphene using a reduction process based on hydrazine (N**2**H**4**) treatment. Per the manufacturer data sheet, the rGO sheets had a conductivity >500 S/m, a diameter 0.5-10 j,m, and a thickness ~1 nm. Before coating, a PDMS fence was created and placed surrounding the Au micropillar array. A 20 iL rGO solution was drop-cast onto the Au micropillar array using a pipette and dried at 80°C for an hour. This process was repeated three times. Though we did not expect the coating to be uniform, the Au pillars were covered by rGO sheets as observed in the SEM images (Figs. 3d, and S1b and S1c of supporting information) and Raman analysis (Fig. 3e). The Au micropillar-rGO surface was further functionalized using SARS-CoV-2 antigens (Fig. 2c). This was achieved by using a coupling reagent consisting of a mixture of EDC (0.2 M) and NHS (0.05 M) in ratios 1:1 by volume. A 20 μL solution of the EDC:NHS mixture was spread over the rGO-Au surface to activate -COOH groups of the rGO sheets. The electrode on the glass substrate was kept in a humid chamber (~100% of humidity) for four hours and washed with PBS solution. Next, a 20 μL of SARS-CoV-2 spike S1 antigen solution (5 μg/mL) was spread on the surface of the rGO-Au array electrode via drop-casting using a pipette (10-50 iL; Cole-Parmer; Vernon Hills, IL 60061) and kept for 4 hr in a humid chamber and then washed with PBS. The activation achieved by EDC:NHS chemistry enabled the primary amine (-NH**2**) groups of the protein molecules (i.e., antigens) of SARS-CoV-2 spike to form C-N bonds with the -COOH groups of rGO sheets via an amidation reaction. In this reaction, the EDC is known to act as a cross-linker and NHS as an activator.^25^ Another SARS-CoV-2 antigen called spike RBD, was also immobilized on a different sensor using same mechanism as that described above. For the RBD antigen, a 20 μL solution (5 μ.g/mL) was used for drop-casting over the Au micropillar-rGO electrode. For all sensors, a 20 iL solution of BSA solution (2 mg/mL) was introduced to the WE surface to block any non-specific sites of the antigen conjugated rGO-Au pillar. In this device, the 3D electrodes are acted as sensitive immunodetectors for COVID 19 antibodies without any labelling agents via antibody-antigen interactions (Fig. 2d).

### Fabrication of PDMS housing and Assembly of the 3DcC Device

The PDMS housing of the 3DcC device depicted was fabricated by soft-replica molding method (Fig. 1e) and integrated with the glass containing functional electrodes (Fig. 1f). A thick polymethylmethacrylate (PMMA) mold was created using high-precision milling machine that had a channel with 1 mm depth, 2 mm and width, and 2 cm length. The channel width in the middle 1 cm section was increased to 2 mm (Fig. 1e, Step 1). A PDMS solution was poured on the PMMA channel to copy an opposite pattern (Fig. 1e, Step2). The bubbles were removed form liquid PDMS mixer by degassing of 1 hour (10^-4^ Torr) in a vacuum chamber. Curing temperature was 80° C for 2 h. The PDMS substrate was peeled off from PMMA channel (Fig 1e, Step 2) and acted a master mold for creating the final microfluidic channel. The surface of this PDMS mold was treated with silicone oil (Ease Release™ 205, Reynoldsam Advanced Materials, Macungie, PA). A new mixer solution of PDMS was then poured into the PDMS mold (Fig. 1e, Step 3) and then peeling off, resulting in the PDMS channel required for the 3DcC device (Fig. 1e, Step 4). Holes were then punched at two ends of the channels by a hollow needle and tygon tubes were inserted for fluid injection. Finally, the PDMS slab was manually placed on the glass substrate such that the CE, WE, and RE were under the channel. Wires were connected to the pads of the electrode outside the PDMS channel using a room temperature curable silver/graphene conductive epoxy to connect to a potentiostat.

### Electrode Characterization

An electrochemical workstation with Zview software (VersaSTAT 3 Potentiostat Galvanostat, Princeton Applied Research, Oak Ridge, TN) were used to record the electrochemical signals and analyze the impedance spectra. In addition, we also enabled a smartphone-based reading platform where the sensor was interfaced with an Android mobile phone using a portable device (Sensit Smart Device, PalmSens, Inc., Randhoeve 221, GA Houten, The Netherlands). The Au micropillar electrodes were imaged by a scanning electron microscope (FEI Sirion SEM, Hillsboro, OR). The elemental analysis was carried out using Energy Dispersive X-ray spectroscopy (EDX) in the same instrument as the SEM imaging. The Raman spectra was collected using the NT-MDT AFM/Raman (NT-MDT America, Tempe, AZ) that used a 532 nm green laser with 12 mW laser power for excitation.

### Electrochemical Simulations

The modeling and 3D simulation of the different electrode structures was conducted using finite element software, COMSOL Multiphysics® (Version 5.5, COMSOL Inc., Burlington, MA). This study was carried out to investigate the diffusion profiles of different geometries of the electrodes and their corresponding electrochemical currents which were generated due to an electrochemical reaction at the surface of the electrode. In this electroanalysis scheme, a redox species B was considered to be oxidized to form a product (A) by losing an electron (B↔A +e^-^). At the boundary, the product concentration is zero, but the bulk concentration of oxidative species is 1 mol/m^3^ and uniform. In this study, Fick’s second law of diffusion was utilized as the domain equation which is given as 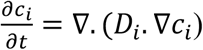, wherein the *c_i_* = 1 mol/m^3^. The diffusivities were taken for the 2D and 3D electrodes as obtained from the cyclic voltammograms at different scan rates, along with the Randles-Sevcik equation (Fig. S3b and S3c).^28^,

## Data Availability

All data needed to evaluate the conclusions in the paper are present in the paper and/or the Supplementary Materials. Additional data related to this paper may be requested from the authors.

## Acknowledgement

The authors would like to acknowledge use of the materials characterization facility such as SEM and Raman at Carnegie Mellon University under grant # MCF-677785.

## Author contributions

R.P. came up with the concept and directed the research. A.A. built the device, developed the various chemistries required for functioning of the device, carried out all the tests, and wrote the first draft of the manuscript. S.J.G. and E. J. provided inputs from biology side and provided several biomolecules used in this research. C.H. and B.Y. carried out the AJ printing of the Au micropillar electrodes. M.S.S. wrote the original program for AJ printing of micropillars. S.J. carried out SEM microscopy, EDX analysis, and Raman studies, along with the simulations of the electrochemical process using COMSOL software. All authors contributed to interpreting the data and preparing and editing the manuscript.

